# A Geographical approach to understand the impacts of Social Determinants of Health on COVID-19 outcomes in an intra-urban context in São Paulo State, Brazil

**DOI:** 10.1101/2025.03.09.25323628

**Authors:** João Pedro Pereira Caetano de Lima, Raul Borges Guimarães

## Abstract

In recent years, health geographers have focused on studies related to COVID-19, proposing frameworks for understanding the Brazilian territory and synthesizing its spatial diffusion pattern. In this process, there is a need for a deeper intra-urban analysis of the impact of COVID-19 on the health of populations in small-to medium-sized cities in non-metropolitan areas. Thus, we aimed to articulate the theory of Social Determinants of Health (SDH) and geoprocessing methodologies to analyze the health situation in the municipality of Presidente Prudente (SP). For this purpose, we used Inverse Distance Weighted (IDW) and Geographically Weighted Regression (GWR) as methodological tools. IDW performs a weighted interpolation among neighbors, generating heat surfaces, whereas GWR executes a spatial regression, presenting correlations between variables. For the analyzed period (March 2020 to July 2021), we found medium-strong correlations between cases and the non-white population, per capita income, and elderly population (R^2^ = 0.46, 0.39, and 0.49, respectively). For deaths, only the elderly population was relevant (R^2^ = 0.34) and the correlation between cases and deaths was satisfactory (R^2^ = 0.65 and 0.51). The results highlight areas of concern, demonstrating that geoprocessing is an important tool for health surveillance, identifying areas and population characteristics that require greater attention, and geography as an important science that helps to understand the impacts of COVID-19.

## 1. Introduction

In recent years, there has been a global effort to understand the impact of COVID-19 worldwide. Within three months of the first recorded case, the disease had reached pandemic proportions and was responsible for reducing global life expectancy by a decade [1]. These data underscore the fact that COVID-19 should be understood as a syndemic, properly located, and contextualized [2, 3].

Given that Brazil ranks 6th in the world in terms of the number of reported infections and 3rd in terms of deaths, it is evident that the disease has severely affected the country’s health, economic, and social systems, creating a serious governance problem [4, 5]. Impacts on the Brazilian population have been identified in the literature through decoding of elementary spatial structures and spatiotemporal patterns of disease diffusion at national and regional scales, contributing to a better understanding of the disease [6, 7].

The same has been worked on for the State of São Paulo, which is marked by being the Brazilian state with the largest population, health equipment, high GDP, and high HDI compared to other Brazilian states. However, intra-urban analyses in small and/or medium-sized cities in non-metropolitan areas lack depth. Thus, we aim to advance the studies we have developed in recent years [8-10], analyzing the intra-urban spatialization of COVID-19 and its possible Social Determinants of Health (SDH) in the municipality of Presidente Prudente (located in the interior of the State of São Paulo). In this way, we assume that no health-disease process is the same among the countries of the world, and therefore, specific studies of the context in Latin America and the Global South are relevant [11-13].

Thus, when seeking relationships between the aforementioned systems and the disease, the theory of SDH becomes relevant, as it predicts the existence of social, environmental, economic, housing, age, race/color, gender, and other factors that influence and/or affect the health status of populations and cause/exacerbate social and health inequalities [14-16]. Therefore, to operationalize the investigation of SDH and its relationship with COVID-19, we discuss the relevance of using geotechnology tools and spatial analysis in medium-sized cities.

Geotechnologies can be understood as the new computational technologies linked to geographical science for mapping information, inserted in the area of Geoprocessing, which uses digital tools to work and process data with spatial characteristics. Spatial analysis is, therefore, the operationalization of this process, allowing the identification and classification of spatial patterns that provide clues for formulating spatial questions [17, 18]. In turn, spatial analysis not only requires the use of techniques but also requires a critical look at this investigative process [19]. Specifically for health issues, these tools have been widely used to investigate patterns of morbidity and mortality, trend analysis, aggravations, among other manifestations of the health-disease process [20-23].

Therefore, we relied on review articles on spatial analysis methodologies and COVID-19 to support our investigation [24-27]. To support the analysis, principles of critical cartography and graphic semiology were incorporated to improve cartographic visualization and effectiveness of mapped information [28-32].

## 2. Materials and Methods

Presidente Prudente is a municipality located in the western region of the State of São Paulo, Brazil, with a population of 225,668 inhabitants. It serves as the regional capital of the Pontal do Paranapanema region, providing a wide range of services including healthcare, commerce, education, leisure, and culture. The municipality has a high Human Development Index (HDI) of 0.806 and a population density of 402.52 inhabitants per km^2^, according to the Brazilian Institute of Geography and Statistics [33]. Due to these characteristics, it is also considered a dispersal hub for COVID-19 in the interior of the São Paulo State and is the headquarters of the 11th Regional Health Department of Sao Paulo (Fig 1).

**Fig 1.**
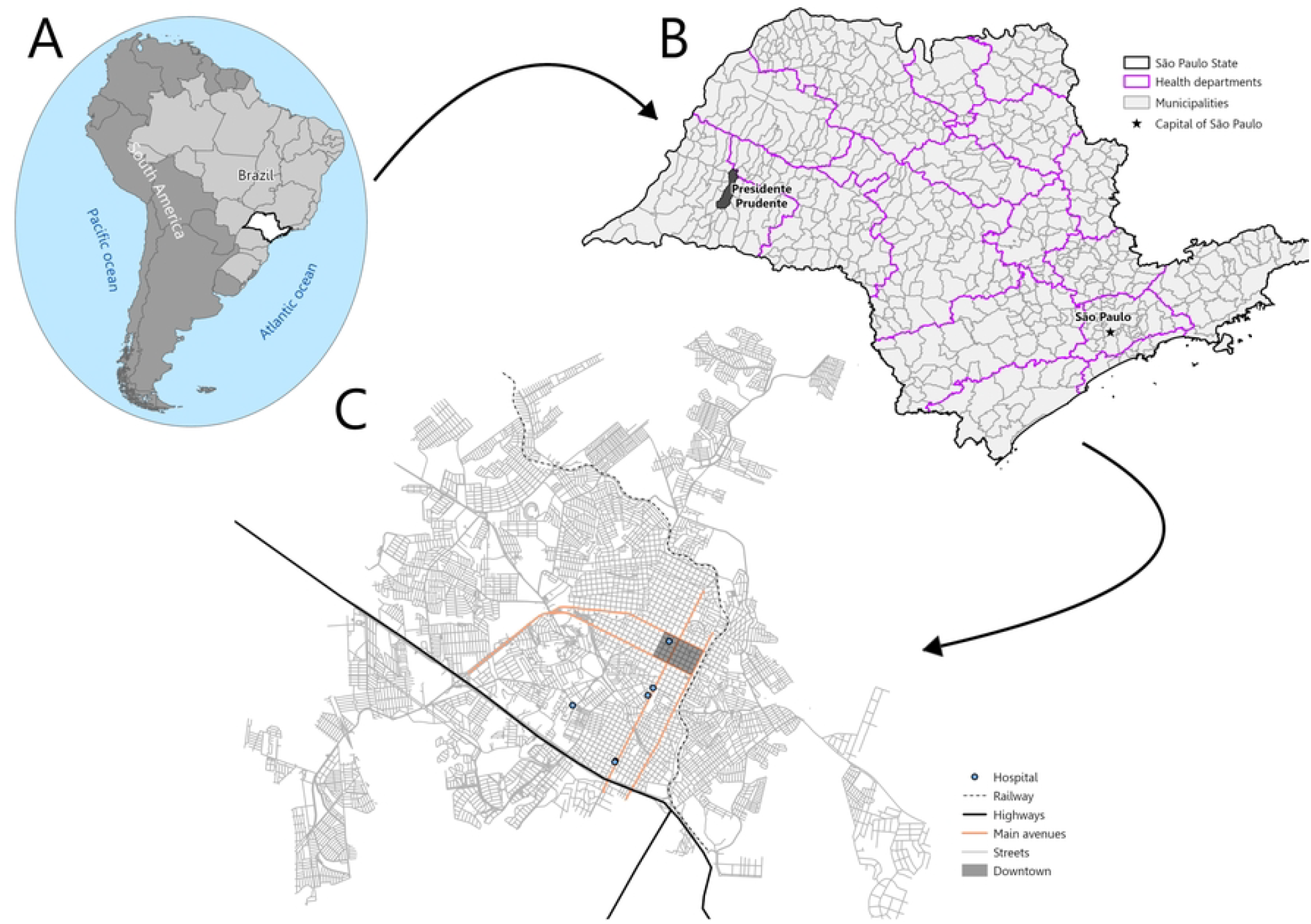
Study area: Presidente Prudente, São Paulo State, Brazil.

**Fig. 2.**
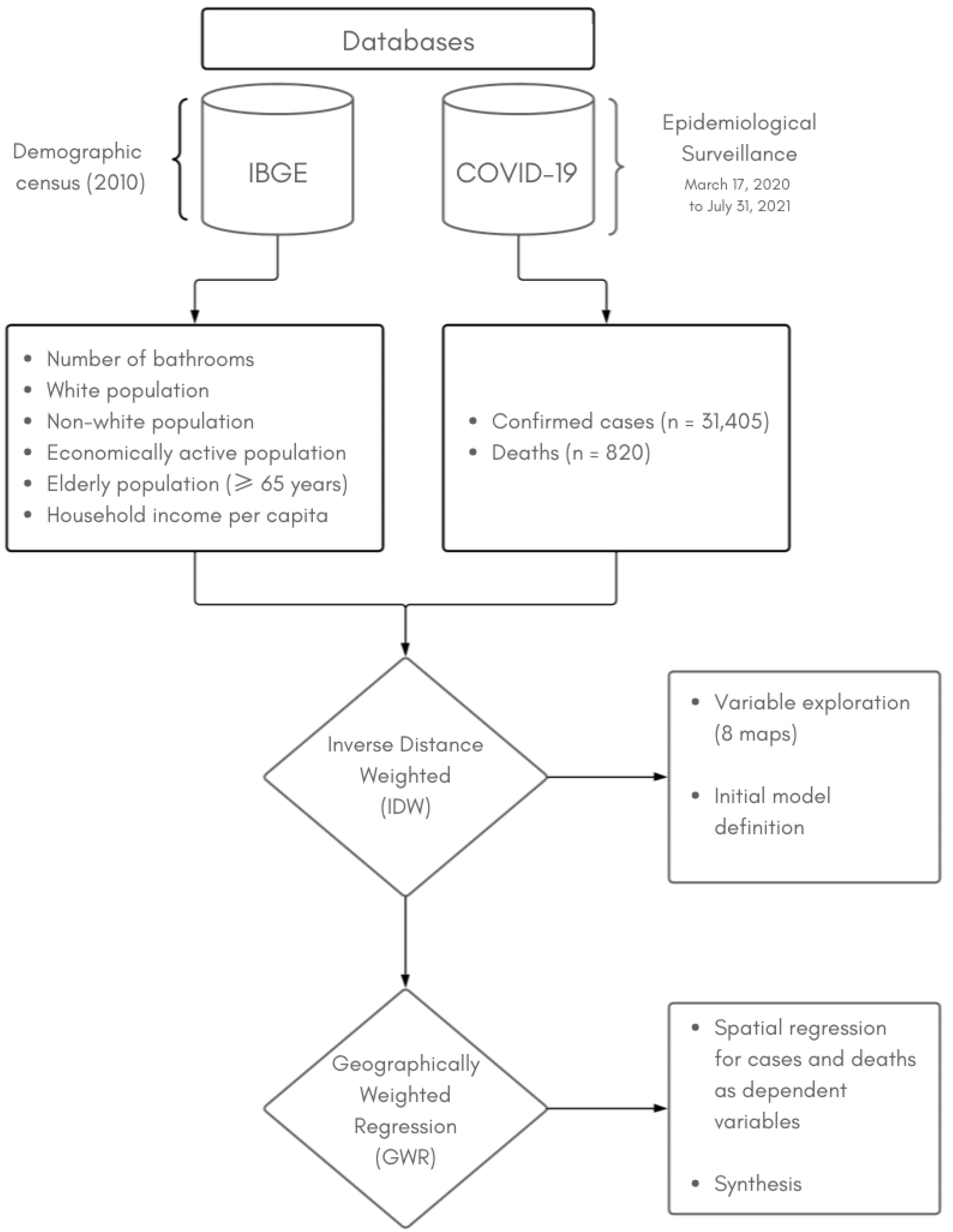
Summary of study design and methodology.

### 2.1. Study design

To conduct this study, we applied different spatial analysis techniques using a logical sequence of data processing and analysis. In an attempt to approximate the SDH to analyze the number of confirmed cases and deaths from COVID-19, we used variables from the last available demographic census and the Epidemiological Surveillance of Presidente Prudente for the period from March 17, 2020, to July 31, 2021 (made available upon request), which marks the intensification of vaccination.

Hence, the total number of confirmed cases in the analyzed period totaled 31,405, and deaths were equivalent to 820. These data were geocoded and mapped using the World Geocoding Service (WGS) in ArcGIS Pro 3.2.2 software [34]. The 2010 census data are made available by the Brazilian Geographic and Statistics Institute based on the SDH previously reported in the literature [35-41]. The variables were i) number of bathrooms in the household, ii) per capita household income, iii) white population, iv) non-white population (summing up people who self-declared as black, brown, and/or indigenous), v) economically active population, and vi) elderly population (≥ 65 years old).

### 2.2. Statistical and spatial analysis

After processing the databases (topological corrections, variable standardization, geocoding, and spatial aggregation), the variables were interpolated using the Inverse Distance Weighted (IDW) method for exploratory spatial data analysis (ESDA) [42]. Geographically Weighted Regression (GWR) was used to correlate confirmed cases and deaths with the previously presented dependent variables and propose the spatial synthesis of the model. It is worth mentioning that these techniques fall within deterministic methodologies, that is, unlike probabilistic ones that estimate values, they consider the exact proximity between events to both interpolate (Fig 3) and correlate (Fig 4) the variables in question [19, 43].

**Fig 3.**
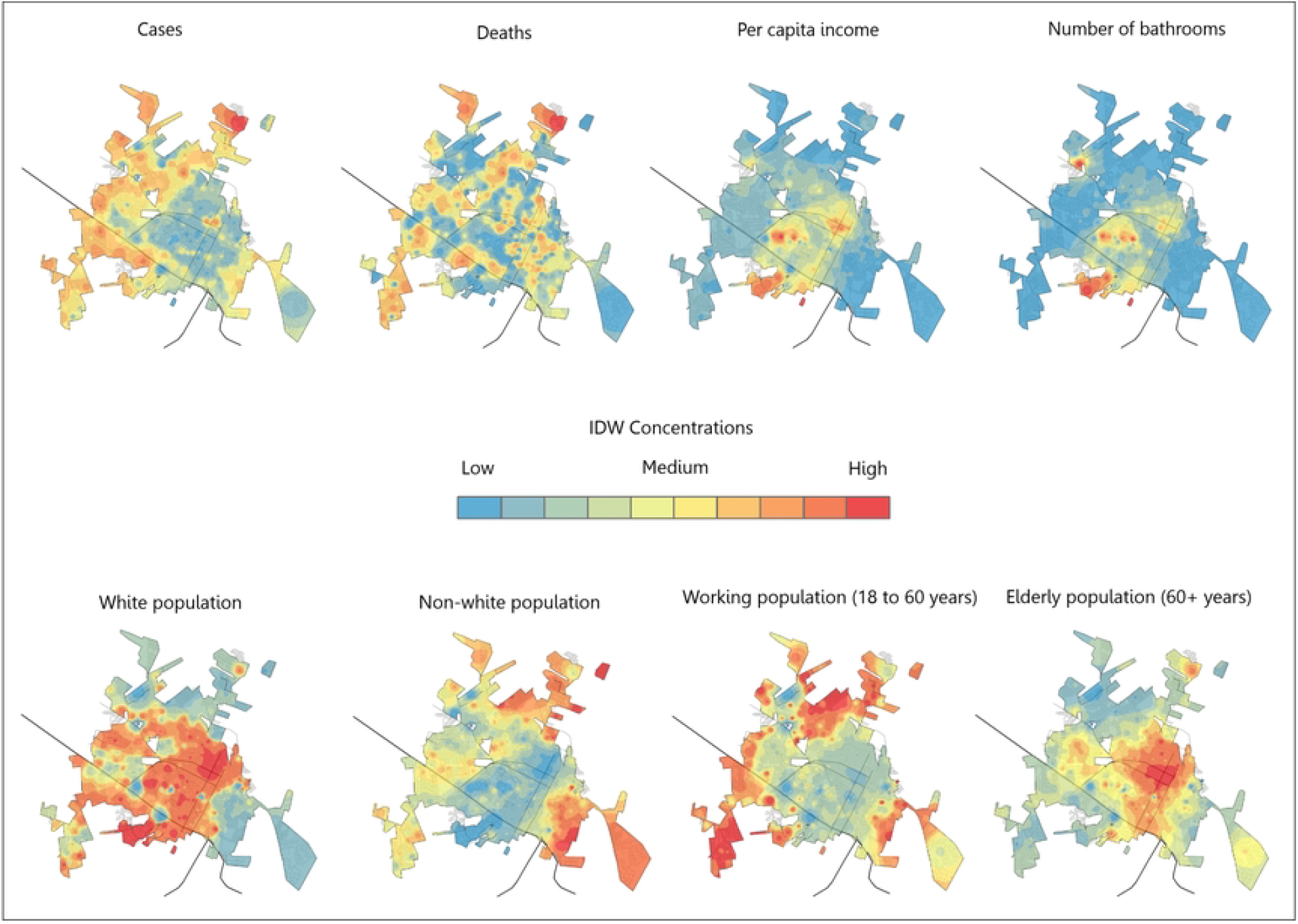
IDW interpolation results for socioeconomic and COVID-19 variables in Presidente Prudente.

**Fig 4:**
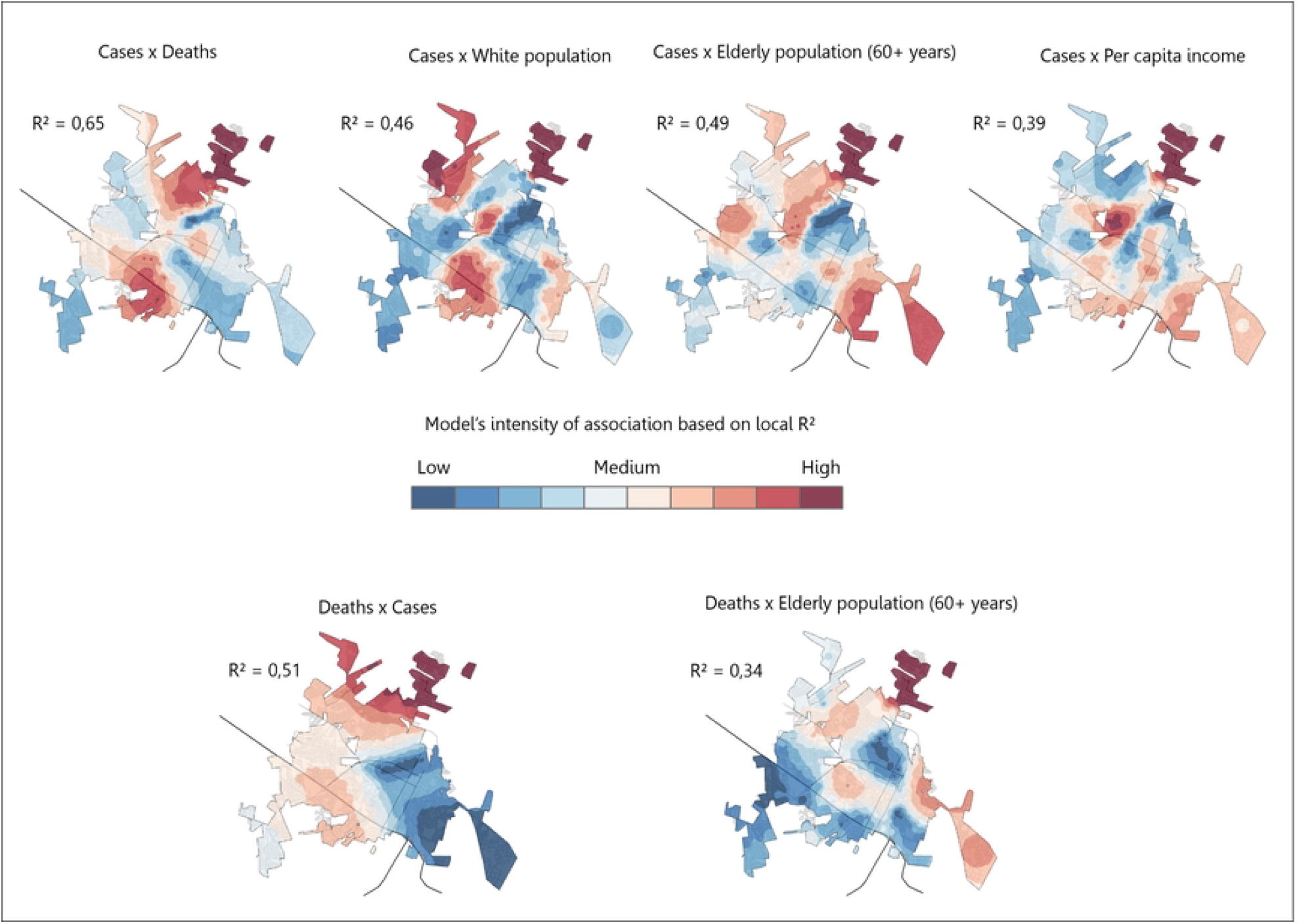
GWR model for confirmed cases and deaths from COVID-19 and sociodemographic variables in Presidente Prudente.

#### 2.2.1. Inverse Distanced Weighted (IDW)

The spatial statistics technique of inverse distance weighting has a long history of use because of its explanatory power in the field of geospatial analysis [44]. Characterized as an exact interpolator, it is used to estimate the concentrations between the values of phenomena that have spatial proximity through a statistical weighting that decays from the spatial distance between events [45]. Thus, it is based on the premise that phenomena that are closer together have more intense relationships with each other and vice versa [46]. Consequently, it returns an estimated surface for the concentration of events. Its scientific notation is as follows:

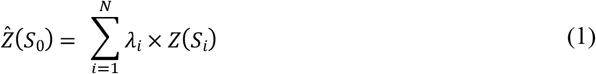

Where 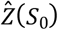 is the value we are predicting for location *S*_0_, *N* is the number of sample points around the prediction location that will be used in the prediction, *λ*_*i*_ are the weights assigned to each measured point that will be used, decreasing with distance, and *Z*(*S*_*i*_) is the observed value of location *S*_*i*_. Finally, the value assigned to the power parameter had a quadratic relationship, and the spatial barriers were the urban census sectors.

#### 2.2.2. Geographical Weighted Regression (GWR)

Geographically Weighted Regression is responsible for translating the statistical regression technique by incorporating the spatial component and local variations into the model, as well as locally assessing the spatial dependence between the selected variables through the R^2^ coefficient, which varies between 0 (null relationship) and 1 (high association). It expresses the coefficient of intensity of the spatial association analyzed, presenting the correlation between the variables [47, 48]. Therefore, it assumes a dependent variable (the one to be explained) and independent variables (which are used to explain the relationship). These characteristics - local analysis, association score, and correlation capacity - configure the utility of the technique for the intra-urban scale analyzed. Its formula is derived from a global regression statistic and can be described as

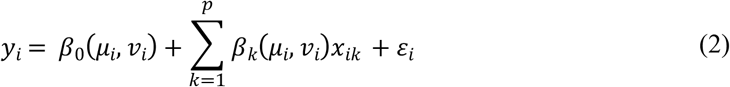

Where *y*_*i*_ represents the observed value, (*μ*_*i*_, *𝒱*_*i*_) are the coordinates of point *i, β*_0_(*μ*_*i*_, *𝒱*_*i*_) is the regression constant *k* of the point *i* and the function of the geographical location of the phenomenon, *p* is the number of independent variables in the model, *x*_*ik*_ corresponds to the value of the independent variable *x*_*k*_ at *i*, and *ε*_*i*_ is the random error coefficient. Finally, the neighborhood method used was the golden search, and the spatial barriers were the urban census sectors.

## 3. Results and discussion

The spatial analysis of sociodemographic variables using IDW revealed distinct spatial distribution patterns, with both direct and inverse relationships between variables (Fig 3). A direct relationship was observed between per capita household income and the number of bathrooms per household, indicating that areas with higher incomes tended to have a greater number of bathrooms. The spatial distribution of the white and non-white populations showed an inverse pattern, with the non-white population predominantly concentrated in peripheral areas characterized by lower income and reduced access to basic services.

Regarding the maps of age patterns in Presidente Prudente, we observed that the economically active population corresponds to the margins of the city, which is also marked by areas of low per capita household income and non-white population, expressing the aforementioned inversely proportional relationship. On the other hand, the elderly population (65 years or older) is concentrated mainly in the central areas of the city.

Sociodemographic variables, such as income, race, and age, are known social determinants of health and are associated with greater vulnerability to COVID-19. The spatial distribution of these variables was highly correlated with the patterns of incidence and mortality from COVID-19 in Presidente Prudente. The concentrations of confirmed cases have a spatial distribution similar to that of the non-white, low per capita income, and economically active populations. This denotes the relationship between the difficulties faced by populations that fall within this social spectrum during the pandemic, that is, the working population of Presidente Prudente, who were often unable to comply with social isolation and lockdown effectively, as they perform essential services in the city’s economy and need to be on the streets during this period. Finally, COVID-19 deaths were concentrated in the northern part of the city, suggesting the presence of local factors that may have contributed to the increase in mortality in this region. Further investigations are needed to identify these factors

Furthermore, the GWR technique allowed us to model spatial relationships in a locally adaptive manner by considering the spatial heterogeneity of the variables. Analyzing the relationship between confirmed cases and deaths, we observed a high association with a coefficient (R^2^) of 0.65, indicating that 65% of the variability in deaths can be explained by confirmed cases, with a higher association in the northern and southern parts of the city, evidencing the spatial heterogeneity of the relationships. In turn, the relationship between cases and the non-white population was considered moderate (R^2^ = 0.46) and locally denoted a higher association in similar portions. When comparing cases with the elderly population (R^2^ = 0.49), we observed a shift in the pattern of local associations to central and eastern areas. Per capita household income was similar in shifting concentrations to central areas and the eastern and southern portions, with a score of R^2^ = 0.39.

Furthermore, we observed that confirmed cases explain the pattern of deaths less intensely than vice-versa, with an R^2^ score of 0.51 (versus 0.65) and a higher association in the northern part of the city (Fig 4). This is relevant because it shows that other variables must be included in the model to better capture the negative outcomes of COVID-19. However, confirmed cases and deaths were relevant pairs. The association between deaths and the elderly population reinforces the importance of social determinants of health, such as age, in identifying vulnerabilities to COVID-19, with a coefficient of 0.39 (R^2^). This association reveals a similar spatial pattern to the previous one, with high associations in the northern portion and medium-high associations in the eastern portion (Fig 4). When analyzing deaths individually, we found in previous studies that most deaths in Presidente Prudente were among the elderly [49].

This initial spatial analysis using GWR provided an overview of the spatial patterns associated with confirmed cases and COVID-19 deaths in the municipality. By combining various sociodemographic and health variables, we were able to identify the areas of greatest vulnerability and direct health surveillance actions.

When modeling confirmed cases, GWR presented a high coefficient of determination (R^2^ = 0.70), indicating that the variables included in the model explained a large part of the spatial variability of the cases. The southern and northern regions had the highest concentrations of cases. Similarly, the combination of confirmed cases and the elderly population to explain deaths resulted in a good model fit (R^2^ = 0.54), with the northern region again showing a critical point.

To identify the areas of greatest concern, we conducted an overlay analysis of high-intensity maps for each model. The intersection of these areas allowed us to delimit regions with the highest risk of COVID-19 transmission and mortality (Fig 5).

**Fig 5.**
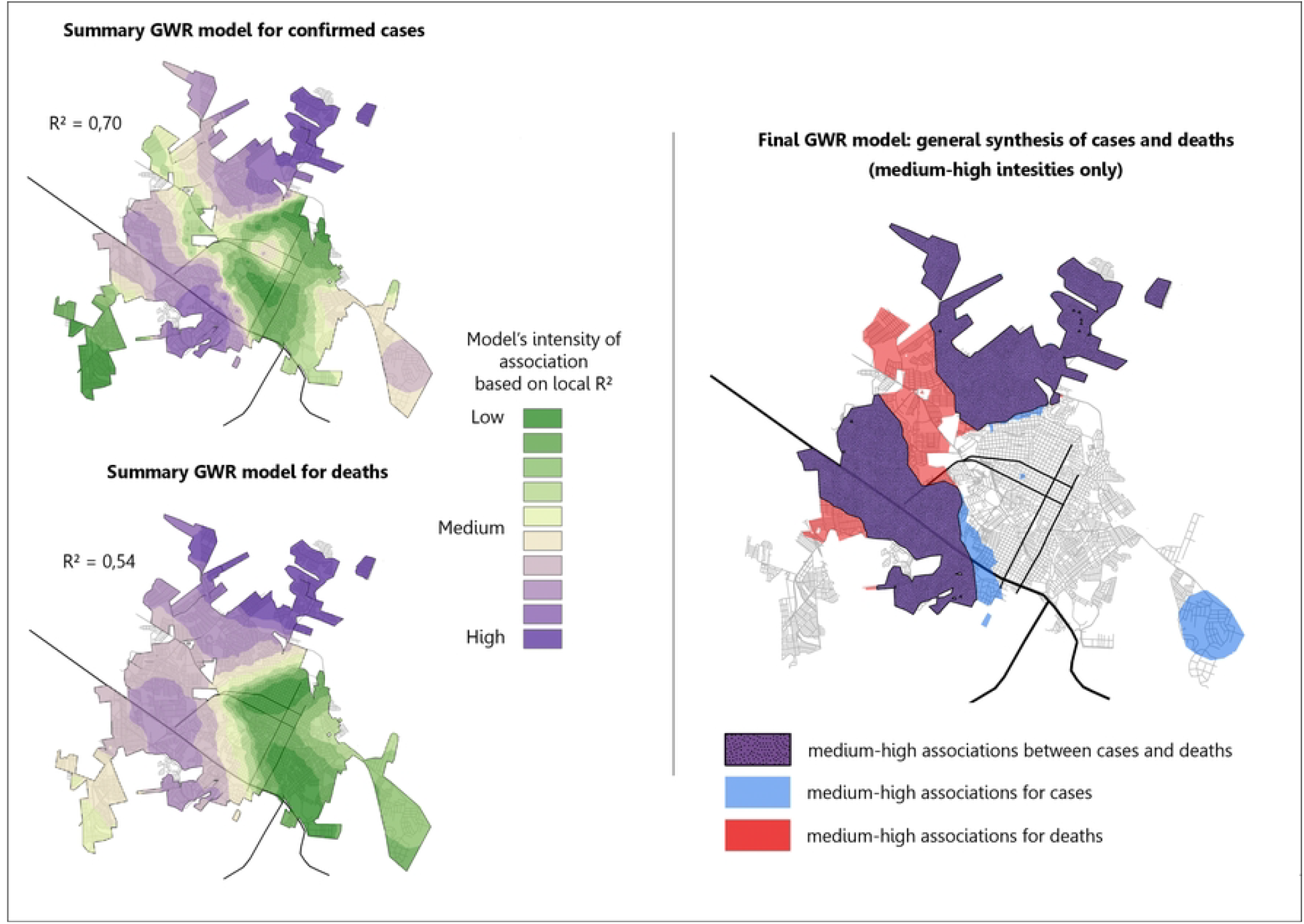
GWR synthesis model for COVID-19 cases and deaths in Presidente Prudente.

## 4. Limitations

It is important to note that GWR, despite its potential, has some limitations, such as the need for a large volume of data and the possibility of multicollinearity among variables. In addition, spatial analysis does not allow the establishment of causal relationships or spatial associations. Another limitation concerns access to sociodemographic data, which was only released in 2010 due to the delay in recensoring and releasing the information by the Brazilian Geographic and Statistics Institute. Thus, we believe that updating the information and deepening the intra-urban analysis of COVID-19 is necessary to advance in the analysis of the impacts on urban communities

## 5. Conclusions

The spatial analysis highlighted areas with the highest vulnerability to COVID-19, which require increased attention from epidemiological surveillance. These areas concentrated on both confirmed cases and deaths, emphasizing the need for specific actions to control the disease in these locations. The analysis also revealed spatial differences in the distribution of cases and deaths, indicating the importance of considering the characteristics of each region when planning intervention strategies.

Hence, the results presented in this study significantly contribute to the understanding of the spatial distribution of COVID-19 in Presidente Prudente, and to the strengthening of epidemiological surveillance in the municipality. The application of spatial analysis techniques, such as IDW and GWR, allows the identification of areas of greater vulnerability and the targeting of disease control and prevention actions.

Although maps are powerful tools for visualizing and communicating results, it is important to recognize that map production involves an interpretive process that can be influenced by social and cultural factors. The subjectivity of both researchers and mapped individuals, as well as existing social structures, can influence how data are collected, analyzed, and cartographically represented [50-52].

Thus, geographic analysis should be understood as part of a broader investigative process that involves the interaction between different disciplines and the consideration of multiple perspectives, demonstrating the importance of a transdisciplinary approach to addressing complex problems, such as the COVID-19 pandemic. The results of this study highlight the crucial role of geography in identifying spatial patterns, understanding the Social Determinants of Health, and formulating more effective public policies for the prevention and control of diseases.

## Data Availability

The socio-economic data comes from the demographic census of the Brazilian Geographic and Statistic Institute, freely accessible at <https://www.ibge.gov.br/en/>. COVID-19 data cannot be shared publicly due to an agreement signed with the Research Ethics Committee, but can be requested through the website: <https://plataformabrasil.saude.gov.br/> Thus, all relevant data are within the manuscript.

## Acknowledgments

We would like to thank the Epidemiological Surveillance Team of Presidente Prudente for their collaboration with the datasets, and the Task Force of Latin American Health Geographers for the discussions in the past years.

## Author Contributions

**Conceptualization:** João Pedro Pereira Caetano de Lima, Raul Borges Guimarães.

**Formal analysis:** João Pedro Pereira Caetano de Lima

**Funding acquisition:** João Pedro Pereira Caetano de Lima, Raul Borges Guimarães.

**Investigation:** João Pedro Pereira Caetano de Lima, Raul Borges Guimarães.

**Methodology:** João Pedro Pereira Caetano de Lima, Raul Borges Guimarães.

**Project administration:** Raul Borges Guimarães

**Software:** João Pedro Pereira Caetano de Lima

**Supervision:** Raul Borges Guimarães

**Visualization:** João Pedro Pereira Caetano de Lima

**Writing – original draft:** João Pedro Pereira Caetano de Lima, Raul Borges Guimarães.

**Writing – review & editing:** João Pedro Pereira Caetano de Lima, Raul Borges Guimarães.

